# Seroprevalence of SARS-CoV-2 in slums and non-slums of Mumbai, India, during June 29-July 19, 2020

**DOI:** 10.1101/2020.08.27.20182741

**Authors:** Anup Malani, Daksha Shah, Gagandeep Kang, Gayatri Nair Lobo, Jayanthi Shastri, Manoj Mohanan, Rajesh Jain, Sachee Agrawal, Sandeep Juneja, Sofia Imad, Ullas Kolthur

## Abstract

**Objective:** Estimate seroprevalence in representative samples from slum and non-slum communities in Mumbai, India, a mega-city in a low or middle-income country and test if prevalence is different in slums.

**Design:** After geographically-spaced community sampling of households, one individual per household was tested for IgG antibodies to SARS-CoV-2 N-protein in a two-week interval.

**Setting:** Slum and non-slum communities in three wards, one each from the three main zones of Mumbai.

**Participants:** Individuals over age 12 who consent to and have no contraindications to venipuncture were eligible. 6,904 participants (4,202 from slums and 2,702 from non-slums) were tested.

**Main outcome measures:** The primary outcomes were the positive test rate for IgG antibodies to the SARS-CoV-2 N-protein by demographic group (age and gender) and location (slums and non-slums). The secondary outcome is seroprevalence at slum and non-slum levels. Sera was tested via chemiluminescence (CLIA) using Abbott Diagnostics Architect^TM^ N-protein based test. Seroprevalence was calculated using weights to match the population distribution by age and gender and accounting for imperfect sensitivity and specificity of the test.

**Results:** The positive test rate was 54.1% (95% CI: 52.7 to 55.6) and 16.1% (95% CI: 14.9 to 17.4) in slums and non-slums, respectively, a difference of 38 percentage points (P < 0.001). Accounting for imperfect accuracy of tests (e.g., sensitivity, 0.90; specificity 1.00), seroprevalence was as high as 58.4% (95% CI: 56.8 to 59.9) and 17.3% (95% CI: 16 to 18.7) in slums and non-slums, respectively.

**Conclusions:** The high seroprevalence in slums implies a moderate infection fatality rate. The stark difference in seroprevalence across slums and non-slums has implications for the efficacy of social distancing, the level of herd immunity, and equity. It underlines the importance of geographic specificity and urban structure in modeling SARS-CoV-2.

---

Investigating the seroprevalence of SARS-CoV-2, and how it varies with population density, is critical for understanding the epidemiology of the disease and tailoring clinical and non-pharmaceutical interventions. Importantly, it informs the vaccination rate required to achieve herd immunity. While there are a growing number of seroprevalence surveys on representative samples^1-3^, there are few published results for low and middle-income countries such as India^4 5^.

India is of particular interest because it has over 2 million confirmed cases, the third largest number worldwide^6^. Moreover, socio-economic disparities and population densities likely impact the distribution of infection in mega-cities like Mumbai. Mumbai, with a population over 12.4 million, has over 135,000 reported cases (as of August 24, 2020)^7^, the most of any Indian city and roughly 5% of confirmed cases in India.

This cross-sectional study estimated seroprevalence in representative samples from slum and non-slum communities in three wards (Matunga, Chembur West, and Dahisar) of Mumbai, one each from the 3 major zones of Mumbai (city, eastern suburbs, and western suburbs, respectively). Our primary outcome is the positive test rate for IgG antibodies to the SARS-CoV-2 nucleocapsid.

## METHODS

Within each of our wards, we recruited subjects separately in areas officially classified as slums and as non-slums. (Slums in this context are communities recognized as living on land to which they do not have legal rights.) Individuals were eligible if they were age 12 years or older and excluded if they refused informed consent or had a contraindication to venipuncture.

### Sample size calculation and study duration

The study was powered to estimate a 1.5 percentage point difference in positive test rate in a two-sided test with 95% confidence in each of the six study areas. Our required sample sizes were 2,249, 1,622, and 564 participants in each of the slum and non-slum sections of Matunga, Chembur West, and Dahisar, respectively, a total of 8,870 individuals.

Because seroprevalence changes over time, we estimate average prevalence over a short, two-week period from June 29 to July 14, 2020, in slums and July 3 to July 19, 2020, in non-slums.

To balance statistical power and bias, we stopped sampling either when we hit sample-size targets or the sampling period lapsed.

### Representative sampling

#### Slums

Within each ward, we recruited in up to 8 of the largest slums by population to balance the fixed costs of working in each additional slum and the possibility that prevalence may vary across slums. We divided each slum into mutually exclusive, geographic polygons covering roughly 400 homes, and sampled 100 homes per polygon. Starting with the home closest to the centroid of each polygon, we sampled one person in every fourth home in one direction.

#### Non-slums

On maps of each ward, we drew rectangular grids such that the ward was covered with just enough cells that, if we draw 100 persons from each cell, we would meet our sample size target for non-slum areas in the ward. We started sampling at a building close to the center of the cell. There were difficulties in obtaining consent from resident associations to enter some buildings to conduct sampling. When allowed to enter, we recruited one household per floor. Otherwise we asked the association to request one volunteer per floor.

Surveyors were given a list of 8 demographic groups (4 age bins by 2 gender bins) and asked to cycle through the list when selecting whom to survey each home. The distribution of our final sample across these groups are a function of the population distribution across groups and by consent rates in each group.

### Data collection, and testing

Each participant was administered a survey to collect socio-demographic data (age, gender, household composition), comorbidities (e.g., hypertension), and contact and travel history over the last 2 months. Phlebotomists collected 5ml of blood from each participant via venipuncture in an EDTA vacutainer. At Kasturba Hospital in Mumbai, plasma was separated and used to test for IgG antibodies via chemiluminescence (CLIA) using Abbott Diagnostics Architect™ N-protein based test. Abbott recommends a 1.4 cutoff for IgG score to label a test result as positive.

### Statistical analysis

We estimated the positive test rate at each of six locations, defined as the slum or non-slum areas in each of 3 wards, in three steps. First, we estimated the positive test rate 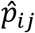 in a demographic group (*i*) in a location *(j)* as the ratio of the number of positive test results and the number of participants that gave an adequate sample. Second, we estimated the positive test rate 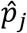 in a location as the weighted average of positive test rates in each demographic group in that location, 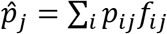, where the weights (Supplement Table e3) are the fraction *f_ij_* of the population in demographic group (*i*) in location *(j)* and ∑*_i_f_ij_ =* 1 Sampling weights were estimated from our survey, which asked how many people in each demographic group reside in a respondent’s household. Third, we aggregate up to the slum or non-slum level across wards with 2011 Census population-weighted averages across wards. The confidence intervals for positive test rates in step one are exact and in steps two and three use normal approximations because we employ weights and location-wise sample sizes are at least 564.

We estimate the seroprevalence of IgG antibodies using the Rogan-Gladen^8^ correction for imperfect accuracy of tests after calculating weighted positive test rates. The estimated sensitivity of CLIA tests range from 90% (95% CI: 74.4% to 96.5%)^9^ to 96.9% (95% CI: 89.5% to 99.5%)^10^, while specificity in those studies was 100% (95% CI: 95.4% to 100%)^9^ and 99.90%^10^, respectively. We present estimates of prevalence assuming both low and high estimates of sensitivity reported and associated estimates of specificity. We employ normal approximations to estimate confidence intervals for prevalence.

### Ethical approval

The study was approved by the IRBs of TIFR (TIFR/IHEC/2020-1), Kasturba Hospital (IRB 20/2020), THSTI (EC/NEW/INST/2019/275), Duke University (Protocol:2020-0575) and University of Chicago (IRB20-1144).

## RESULTS

We analyzed 4,202 samples from slums and 2,702 from non-slums. Supplement Table e1 reports the number of individuals with test results in each demographic group by location. We did not achieve sample size targets in two weeks in 2 non-slum areas due to low consent rates, driven primarily by fear of getting infected during testing. Supplement Table e2 presents demographic information on participants by community.

Figure 1 provides estimates of positive test rates by age and gender groups in different communities. Table 1 reports that positive test rates of 54.1% and 16.1% in slums and in non-slums, respectively. The difference of 38.0 percentage points is highly significant (p<0.001). Underlying IgG scores are likewise higher in slums than non-slums (Supplement Figure e1). The positive test rates are more sensitive to the manufacturer’s recommended cutoff for positive tests in slums than in non-slums (Supplement Figure e2).

**Figure 1.**
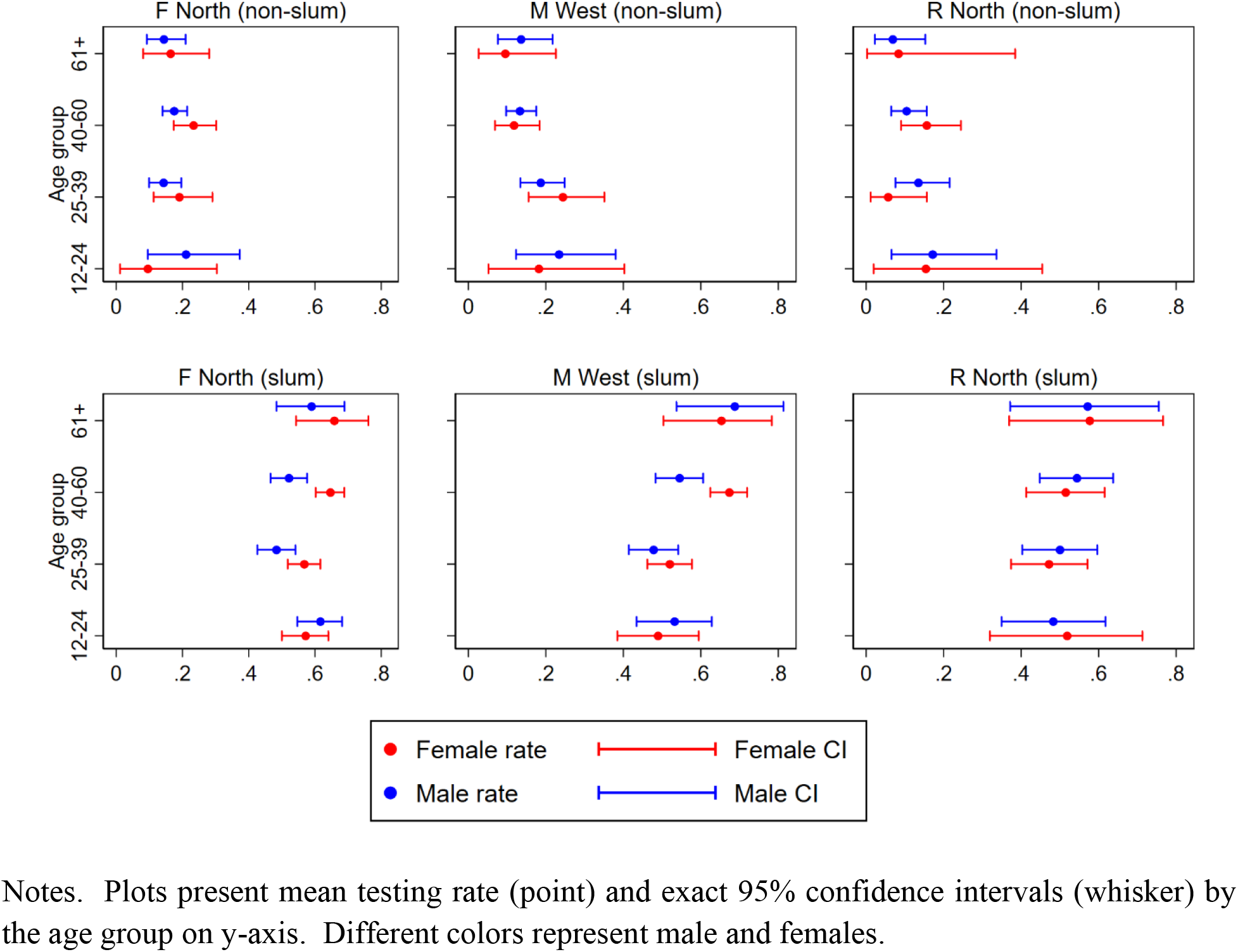
Positive test rate by age group and sex for each location.

**Table 1.** Positive test rate and prevalence, by location and estimate of test accuracy

| Location | Positive test |  | Prevalence |  |  |  |
| --- | --- | --- | --- | --- | --- | --- |
|  |  |  | (sensitivity = 0.969,<br>specificity = 0.999) |  | (sensitivity = 0.900,<br>specificity = 1.000) |  |
|  | Rate | 95% CI | Rate | 95% CI | Rate | 95% CI |
| Non-slums | 0.161 | (.149,.174) | 0.164 | (.151,.176) | 0.173 | (.16,.187) |
| Matunga | 0.179 | (.158,.199) | 0.181 | (.16,.202) | 0.192 | (.17,.214) |
| Chembur West | 0.167 | (.145,.189) | 0.169 | (.147,.192) | 0.179 | (.156,.203) |
| Dahisar | 0.119 | (.095,.143) | 0.120 | (.095,.145) | 0.128 | (.102,.154) |
| Slum | 0.541 | (.527,.556) | 0.558 | (.543,.572) | 0.584 | (.568,.599) |
| Matunga | 0.572 | (.553,.592) | 0.590 | (.569,.61) | 0.617 | (.596,.638) |
| Chembur West | 0.551 | (.528,.574) | 0.568 | (.544,.591) | 0.594 | (.57,.618) |
| Dahisar | 0.510 | (.471,.548) | 0.525 | (.485,.565) | 0.549 | (.507,.591) |

Note. The positive test rate is estimated by, first, estimating the rate for each demographic group and, then, calculating a weighted average that ensures each group's rate has a weight proportional to its share of the population in a location, with weights as indicated in Table e3. Prevalence and its confidence interval is calculated from the weighted average positive test rate for a location using the Rogan-Gladden formula<sup>8</sup>. Confidence interval is estimated using a normal approximation.

Accounting for test sensitivity and specificity, seroprevalence is also higher in slums, with means ranging from 55.8% to 58.4% in slums and 16.4% to 17.3% in non-slums across locations (Table 1).

## DISCUSSION

Our findings imply high seroprevalence in the slums we surveyed. The epidemic may be in advanced stages in those locations, perhaps due to higher density in slums^11^. The high seroprevalence rate raises the possibility that a high fraction of cases is asymptomatic and has implications for the efficacy of social distancing policies in slums. The implied past infection rate during the 4 months after the first SARS-CoV-2 case in Mumbai, reported on March 12, 2020, suggests a high reproductive rate in slums.

Slums constitute 49% of the population (slum population 705,523; non-slum population 709,394) of the 3 wards in our sample according to the city’s Mid Year Estimated Population for 2019^12^. The Brihanmumbai Municipal Corporation reported 292 and 203 deaths as July 13 and July 20, dates closest to the last day of sampling, in slum and non-slum, respectively, in these wards. Taken together our estimate of seroprevalence, these number imply an infection fatality rate of 0.076% and 0.263% in non-slums. The overall rate is estimated to be 0.12%.

Our findings also suggest that seroprevalence is more than 3 times higher in slums than non-slum areas in the same ward. The difference highlights the importance of geographic specificity in modeling^13^ and modeling variation in contact rates^14^. The implied variation in reproductive rates also has implications for the level of herd immunity city-wide^14^. This in turn informs estimates of the vaccination rate required to reach that level. Differences in income and health between slums and non-slums^15^ suggest that the epidemic may have significant implications for equity.

Our study generates novel hypotheses such as gender and age-specific variations in exposure to SARS-CoV2 and subsequent immune response. The stark difference in prevalence between slums and non-slums highlights the effect of crowding on infection spread. Combining these insights will be critical to estimate the herd immunity level in settings with high intermixing of populations with different demographics and urban infrastructure.

Overall, this study will likely have implications on our efforts to tackle SARS-CoV-2 in mega-cities and in low- and middle-income countries.

## Data Availability

Data availability remains under consideration.

## ACKNOWLEDGEMENTS

The study was sponsored by Action Covid19 Team, A.T.E. Chandra Foundation, and Godrej Industries.

We thank the Municipal Corporation of Greater Mumbai (Mr. Suresh Kakani, Dr. Mangla Gomare, the Assistant Commissioners and Municipal Officers of Health of the three wards) for their support during the duration of the project. The authors also thank Ms. Poonam Choksi (ATE Chandra Foundation), Ms. Priya Maisheri (CDSA), and Mr. Sandeep Singhal (Nexus Venture Partners) for their contribution to the study. Finally, we thank all field teams and participants involved in this project.

## ONLINE SUPPLEMENT

Table e1 provides data on the breakdown of our sample by age group, gender, and location, where location is defined both by ward and whether the community is a slum or not. Table e2 provides additional demographic detail by aggregating, without weights, across slums and across non-slums. Our sample has fewer females in non-slums. It has members in the youngest age group (12-24 years old) and fewer in the oldest age group (61 and older) in slums. The age distribution in our sample is consistent with that in the general population (Table e3), which is younger in slums. We use the age distribution of the slum and non-slum population in Table e3 as weights to convert positive test rates at the demographic group and location level to positive test rates at the location level.

We examine the sensitivity of our estimates of positive test rates to the cutoff of IgG titer used to label test results as positive or negative. The manufacturer recommends a cutoff of 1.4 for its Architect CLIA test for N-antibodies to SARS-CoV-2. Figure e1 shows that the cumulative distribution of IgG titers in slums stochastically dominates that in non-slums, i.e., average titer is higher in slums. Non-slums have a much higher concentration (roughly 75% of the population) than slums (roughly 25%) of IgG concentrations near zero. While there is no jumps in the data around the 1.4 cutoff, changes in the cutoff impact estimates of the positive rate in slums much more than non-slums (Table e5).

**Table e1.**
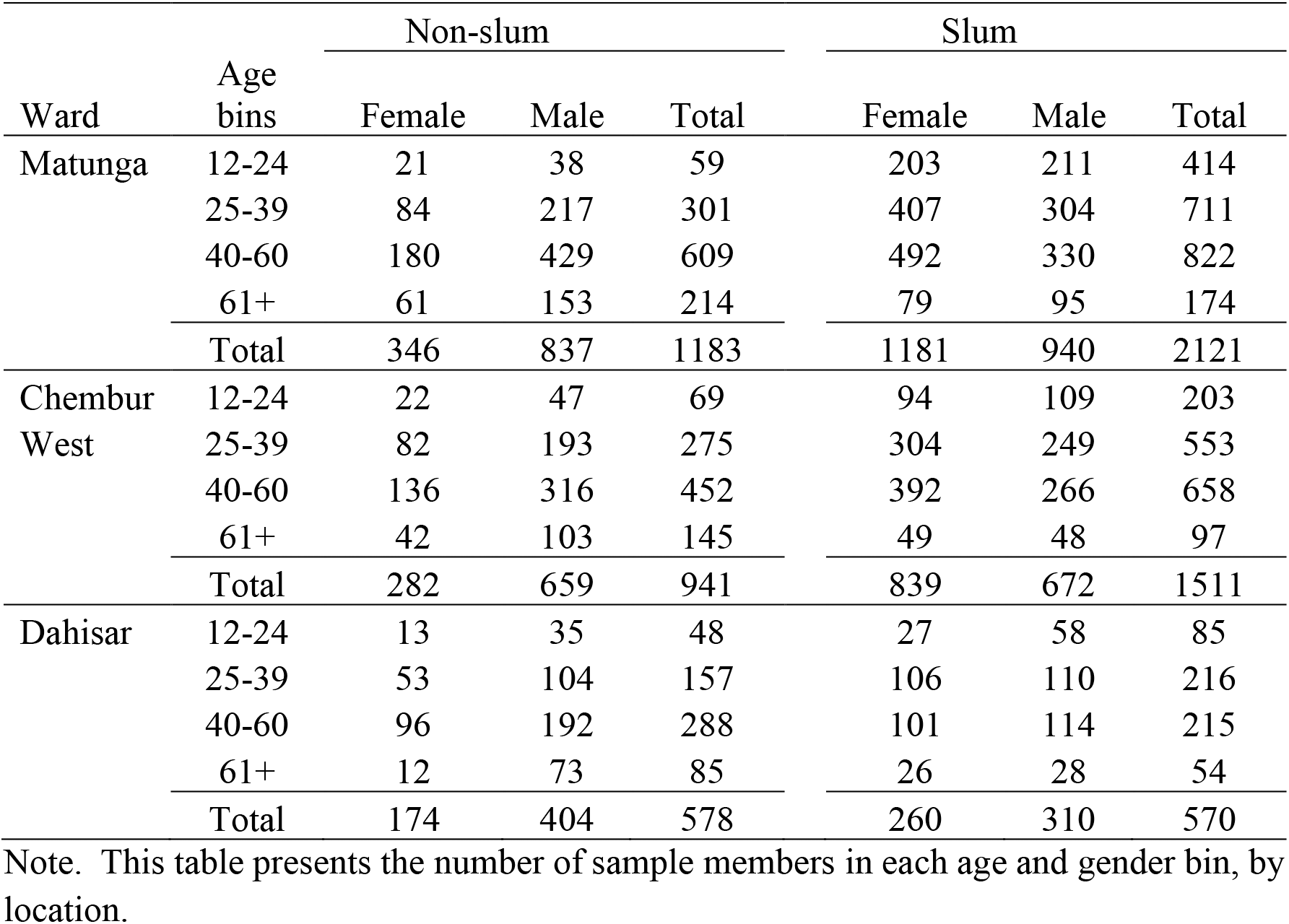
Demographic profile of sample, by location.

| Ward | Age bins | Non-slum |  |  | Slum |  |  |
| --- | --- | --- | --- | --- | --- | --- | --- |
|  |  | Female | Male | Total | Female | Male | Total |
| Matunga | 12-24 | 21 | 38 | 59 | 203 | 211 | 414 |
|  | 25-39 | 84 | 217 | 301 | 407 | 304 | 711 |
|  | 40-60 | 180 | 429 | 609 | 492 | 330 | 822 |
|  | 61+ | 61 | 153 | 214 | 79 | 95 | 174 |
|  | Total | 346 | 837 | 1183 | 1181 | 940 | 2121 |
| Chembur West | 12-24 | 22 | 47 | 69 | 94 | 109 | 203 |
|  | 25-39 | 82 | 193 | 275 | 304 | 249 | 553 |
|  | 40-60 | 136 | 316 | 452 | 392 | 266 | 658 |
|  | 61+ | 42 | 103 | 145 | 49 | 48 | 97 |
|  | Total | 282 | 659 | 941 | 839 | 672 | 1511 |
| Dahisar | 12-24 | 13 | 35 | 48 | 27 | 58 | 85 |
|  | 25-39 | 53 | 104 | 157 | 106 | 110 | 216 |
|  | 40-60 | 96 | 192 | 288 | 101 | 114 | 215 |
|  | 61+ | 12 | 73 | 85 | 26 | 28 | 54 |
|  | Total | 174 | 404 | 578 | 260 | 310 | 570 |

Note. This table presents the number of sample members in each age and gender bin, by location.

**Table e2.** Demographic features of the sample, unweighted and by residence in slum

|  | Count | Mean | (Exact 95% Conf. Int.) |  |
| --- | --- | --- | --- | --- |
| Non-slum | 2702 |  |  |  |
| Female | 802 | 0.297 | 0.280 | 0.314 |
| Ages 12-24 | 176 | 0.065 | 0.056 | 0.075 |
| Ages 25-39 | 733 | 0.271 | 0.255 | 0.288 |
| Ages 40-60 | 1349 | 0.499 | 0.480 | 0.518 |
| Ages 61+ | 444 | 0.164 | 0.151 | 0.179 |
| Slum | 4202 |  |  |  |
| Female | 2280 | 0.543 | 0.527 | 0.558 |
| Ages 12-24 | 702 | 0.167 | 0.156 | 0.179 |
| Ages 25-39 | 1480 | 0.352 | 0.338 | 0.367 |
| Ages 40-60 | 1695 | 0.403 | 0.388 | 0.418 |
| Ages 61+ | 325 | 0.077 | 0.069 | 0.086 |

Note. Table presents the fraction of slum and non-slum population that are female and in each age group.

**Table e3.** Age distribution among the population in each community, by sex, ward and slum status

| Ward | Age group | Non-slums |  |  | Slums |  |  |
| --- | --- | --- | --- | --- | --- | --- | --- |
|  |  | No of hhd members | Male (share) | Female (share) | No of hhd members | Male (share) | Female (share) |
| Matunga | All | 3487 | 0.546 | 0.454 | 8936 | 0.505 | 0.495 |
|  | 12-24 | 626 | 0.102 | 0.078 | 2916 | 0.175 | 0.151 |
|  | 25-39 | 947 | 0.144 | 0.127 | 3201 | 0.176 | 0.182 |
|  | 40-60 | 1311 | 0.208 | 0.168 | 2293 | 0.123 | 0.133 |
|  | 61+ | 603 | 0.092 | 0.081 | 526 | 0.030 | 0.029 |
| Chembur West | All | 3925 | 0.521 | 0.479 | 6433 | 0.502 | 0.498 |
|  | 12-24 | 846 | 0.119 | 0.096 | 1752 | 0.146 | 0.127 |
|  | 25-39 | 1078 | 0.140 | 0.135 | 2206 | 0.173 | 0.170 |
|  | 40-60 | 1456 | 0.191 | 0.180 | 2057 | 0.153 | 0.167 |
|  | 61+ | 545 | 0.070 | 0.069 | 418 | 0.030 | 0.035 |
| Dahisar | All | 5004 | 0.522 | 0.478 | 3386 | 0.545 | 0.455 |
|  | 12-24 | 1069 | 0.109 | 0.105 | 1088 | 0.190 | 0.131 |
|  | 25-39 | 1389 | 0.143 | 0.134 | 1055 | 0.160 | 0.152 |
|  | 40-60 | 1792 | 0.189 | 0.169 | 906 | 0.141 | 0.127 |
|  | 61+ | 754 | 0.081 | 0.070 | 337 | 0.054 | 0.045 |

Note. These age distributions are calculated from surveys of sample households. The surveys asked the number of individuals in age group by gender in the sample member's household. The fractions sum to one within a location. Calculations drop observations where sample member does not answer question about household composition.

**Figure e1.**
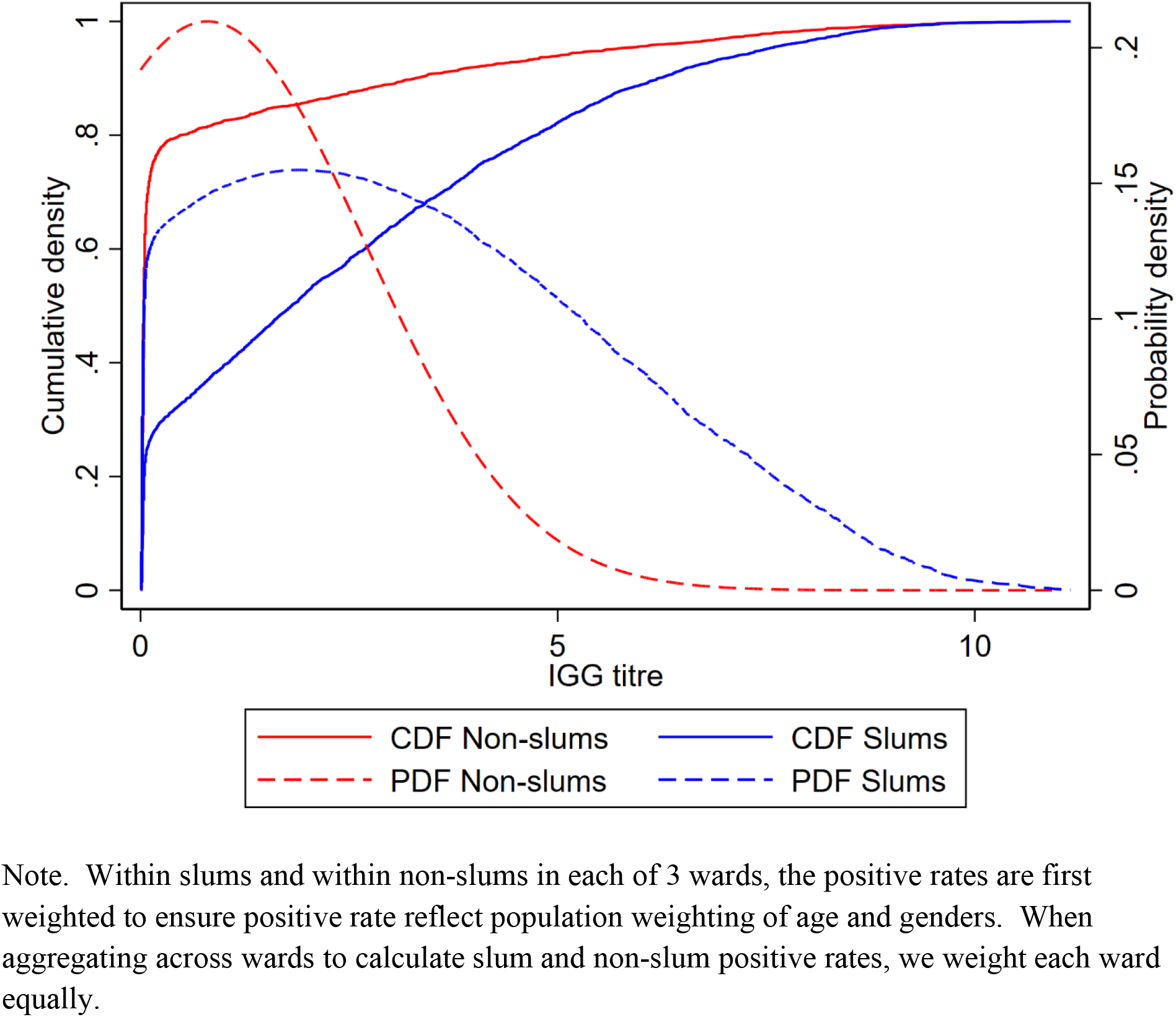
Empirical cumulative and probability density functions for IGG scores, separately for slum and non-slum communities and weighted to reflect age and gender distribution in the population.

**Figure e2.**
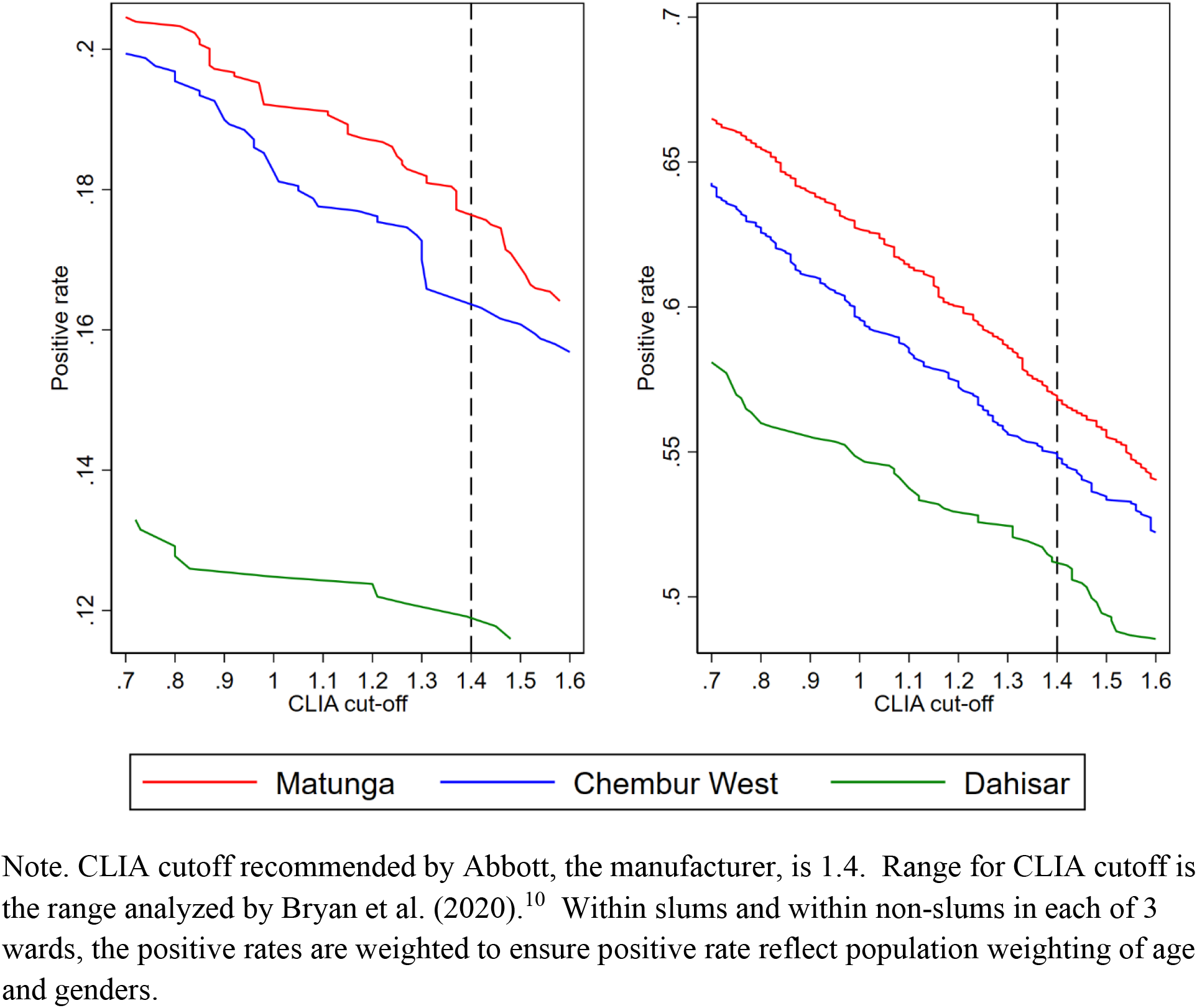
Relationship between weighted positive test rate and CLIA cut-off value, by ward for non-slums and slums

